# Reconstruction of SARS-CoV-2 transmissibility within households in the UK Virus Watch Study

**DOI:** 10.1101/2025.06.12.25329406

**Authors:** Laura Buggiotti, Arturo Torres Ortiz, Xavier Didelot, Alexei Yavlinsky, Cyril Geismar, Jan Kovar, Charles Miller, Luz Marina Martin Bernal, Rachel Williams, Ibrahim Abubakar, Robert W. Aldridge, Judith Breuer

**Affiliations:** Infection, Immunity and Inflammation, Great Ormond Street Institute of Child Health, University College London, London, UK; School of Life Sciences and Department of Statistics, University of Warwick, Coventry, UK; Institute of Health Informatics, University College London, London, UK; Institute of Epidemiology and Health Care, University College London, London, UK; Great Ormond Street Hospital for Children NHS Trust, London, UK; Genetics and Genomic Medicine Department, Great Ormond Street Institute of Child Health, University College London, London, UK; Faculty of Population Health Sciences, University College London, London, UK; Centre for Public Health Data Science, Institute of Health Informatics, University College London, London, UK

## Abstract

Households provide ideal settings to study SARS-CoV-2 transmission due to close proximity and extended exposure among members. Understanding transmission patterns is crucial for implementing effective control measures. We analysed whole genome sequences from 237 subjects across 162 households within the Virus Watch Prospective Community Cohort Study (372 total participants). We incorporated minority variants as indicators of within-host genomic diversity into phylogenetic models to reconstruct viral relationships more accurately than using consensus sequences alone. In 73/162 households with multiple infections, phylogeny identified eight secondary cases resulting from separate introduction events rather than within-household transmission. For the remaining 65 households, including minority variants resolved transmission chains that were otherwise uncertain. Our approach demonstrates that integrating within-host genetic diversity into phylogenetic models alongside epidemiological data provides a robust method to accurately identify household transmission events, assess infection risk factors, and improve secondary attack rate estimation, essential for understanding viral spread dynamics.

## Introduction

Households represent ideal settings to study the mode of viral transmission of SARS-CoV-2 given the proximity and duration of exposure. Studying transmission in this environment offers unique insights into how the virus spreads, evolves, and adapts^1–6^. Deep whole-genome sequencing of SARS-CoV-2 from household members enables the identification of viral variants, allowing exploration of the dynamics of viral transmission and clarifying whether transmission occurred exclusively within the household or if multiple introductions took place instead.

The integration of genomics and phylogenetics together with epidemiology in household transmission studies provides a powerful framework for understanding the intricate dynamics of viral spread in close-contact settings. In fact, the granularity of the sequence data and the amount of genomic diversity represent key factors to infer transmission. To date, the relatedness of viral sequences identified from potential transmission chains versus community virologic surveillance has been compared using phylogenetic trees of whole-genome consensus sequences^5,7^. However, the slow mutation rate of SARS-CoV-2^8^ and the lack of genetic differences that can occur between unrelated samples, particularly following recent emergence of a new variant, might result in poor/absent phylogenetic signal with unrelated viruses within the household apparently sharing the same consensus sequence^9^. To increase the genetic resolution of transmission inference, some authors have proposed leveraging within-host diversity by capturing minority variants alongside consensus sequences. The inclusion of minority variants, which may emerge and fluctuate rapidly within hosts, enhance the chances of detecting distinguishing mutations among individuals within the same household^10,11^.

Transmission inference using genetic variants can be complicated by the size of the transmission bottleneck, as only a subset of viral diversity is successfully transmitted between hosts, potentially leading to misclassification of transmission links^12^. In an effort to further understand the dynamics of transmission, its bottleneck was also investigated. Some studies point to a narrow transmission bottleneck^8,13,14^ in SARS-CoV-2 that significantly reduces viral genetic diversity at the start of each infection. However, it might be also possible that the transmission of viral genetic diversity could show different patterns in different settings, such as superspreading events^15,16^ or a more conducive environment, as in the case of a household.

In this study we used whole-genome sequencing of SARS-CoV-2 from a subset of participants of Virus Watch^17^, a prospective community cohort study in England (participants recruited between June 2020 and March 2022). We took advantage of the within-host genomic diversity approach developed by Torres Ortiz *et al*. 2023^11^ in which low frequency within samples variation were included in a phylogenetic model to reconstruct the phylogenetic relationship, leading to more accurate transmission dynamics than if only the consensus variation was used. This method assumes a wider transmission bottleneck than has previously been described^8,13,14^, a feature which we examine further.

## Results

### Study participants

A total of 162 households, comprising 372 participants, with at least one positive participant, from the Virus Watch prospective community cohort study^17^ were analysed (Table 1, Supplementary Figure 2).

**Table 1.** Household characteristics.

| **Age (years)** | **n** | **%** |
| --- | --- | --- |
| 3-24 | 54 | 14.63 |
| 25-34 | 11 | 2.98 |
| 35-44 | 25 | 6.78 |
| 45-54 | 55 | 14.91 |
| 55-64 | 99 | 26.83 |
| 65-74 | 101 | 27.37 |
| 75+ | 24 | 6.50 |
| **Household size** |  |  |
| 1 | 16 | 9.88 |
| 2 | 106 | 65.43 |
| 3 | 19 | 11.73 |
| 4 | 18 | 11.11 |
| 5 | 3 | 1.85 |
| **Vax/participant/HH** |  |  |
| 0 | 9 | 2.42 |
| 1 | 12 | 3.23 |
| 2 | 53 | 14.25 |
| 3 | 62 | 16.67 |
| 4 | 108 | 29.03 |
| 5 | 90 | 24.19 |
| 6 | 19 | 5.11 |
| 7 | 16 | 4.30 |
| 8 | 3 | 0.81 |

The number of participants per household varied between one and five with the majority having two (65.4 %) and the age of participants were between 3 and 94 years. In 73/162 (45.1%) households more than one participant became infected following an index case.

Samples were collected between 20/12/2022 and 30/06/2023 when various Omicron subvariants were circulating in the UK. A total of 336 participants sent lateral flow positive samples, of which 240 were above the viral load threshold for whole genome sequencing and 237 passed the quality control threshold (>90% genome coverage at 10X sequencing depth) and were further analysed. As expected, all the participants were positive for the Omicron variant, with Pangolin^21^ assigning the majority (69.6%) of genomes as BA.2-like, and XBB.1.5 identified as the predominant subvariant (40.5%; Figure 1). To compare the lineage distributions observed in our cohort with broader population trends, we analysed contemporaneous UK-wide genomic surveillance data (Supplementary Figure 3). While XBB.1.5 was the predominant lineage across the UK during the study period, the genomic Virus Watch cohort exhibited a higher proportion of BA.2-like genomes during May–June, a trend not observed in the general population. This discrepancy suggests potential differences in exposure patterns, transmission dynamics, or cohort characteristics that may not be fully captured by national surveillance data.

**Figure 1.**
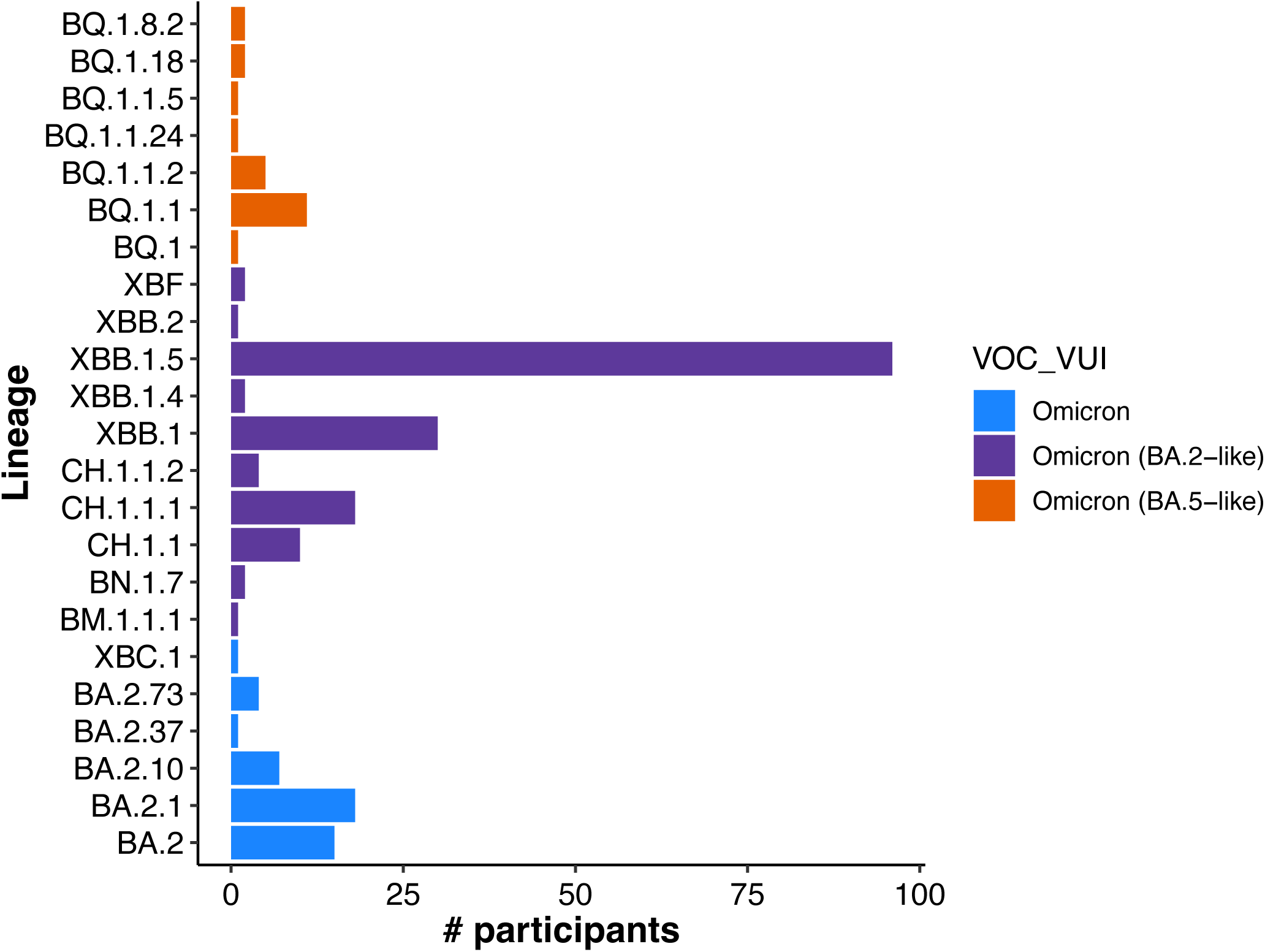
Lineage assignation

### Phylogenetic analysis

Both consensus and within-host diversity maximum likelihood phylogenetic trees were built. The latter uses a model of nucleotide evolution with 16-character states accounting for within-sample variation which is coded in the genome alignment using existing IUPAC nomenclature, as previously described^11^. Pairwise branch distances between SARS-CoV-2 positive samples were significantly higher in participants belonging to different households compared to those coming from the same household in both consensus and within-host diversity phylogenetic trees. Within-host diversity phylogenetic trees resulted in greater branch distances thereby increasing the distinction between household transmission pairs as compared with sequences from unrelated individuals in different households (Figure 2). The within-host genomic diversity analysis, which incorporates minority variants in phylogenetic reconstruction, confirmed 64 transmission pairs which had previously been identified by consensus sequence phylogeny and identified an additional pair that had been poorly resolved using consensus methods (Figure 3, Supplementary Figure 4). In 73 out of 162 households, more than one participant became infected following an index case. Both consensus and minority variant phylogenetic trees identified separate introduction events rather than within-household transmission in eight households. In four of these cases, the index and recipient individuals carried different Omicron lineages. In the remaining four households, although the Omicron lineage was the same, the index and household contact sequences were clearly separated in the phylogenetic tree and did not cluster closely with either the index case or cases from other households. An additional 4,219 high-coverage consensus sequences collected in the UK over the six-month study period were analysed alongside genomic data from the Virus Watch study to assess whether inferred transmission events remained clustered. The phylogenetic tree generated using this combined dataset confirmed that individuals from the same household continued to cluster together, as previously observed at the consensus sequence level (Supplementary Figure 5).

**Figure 2.**
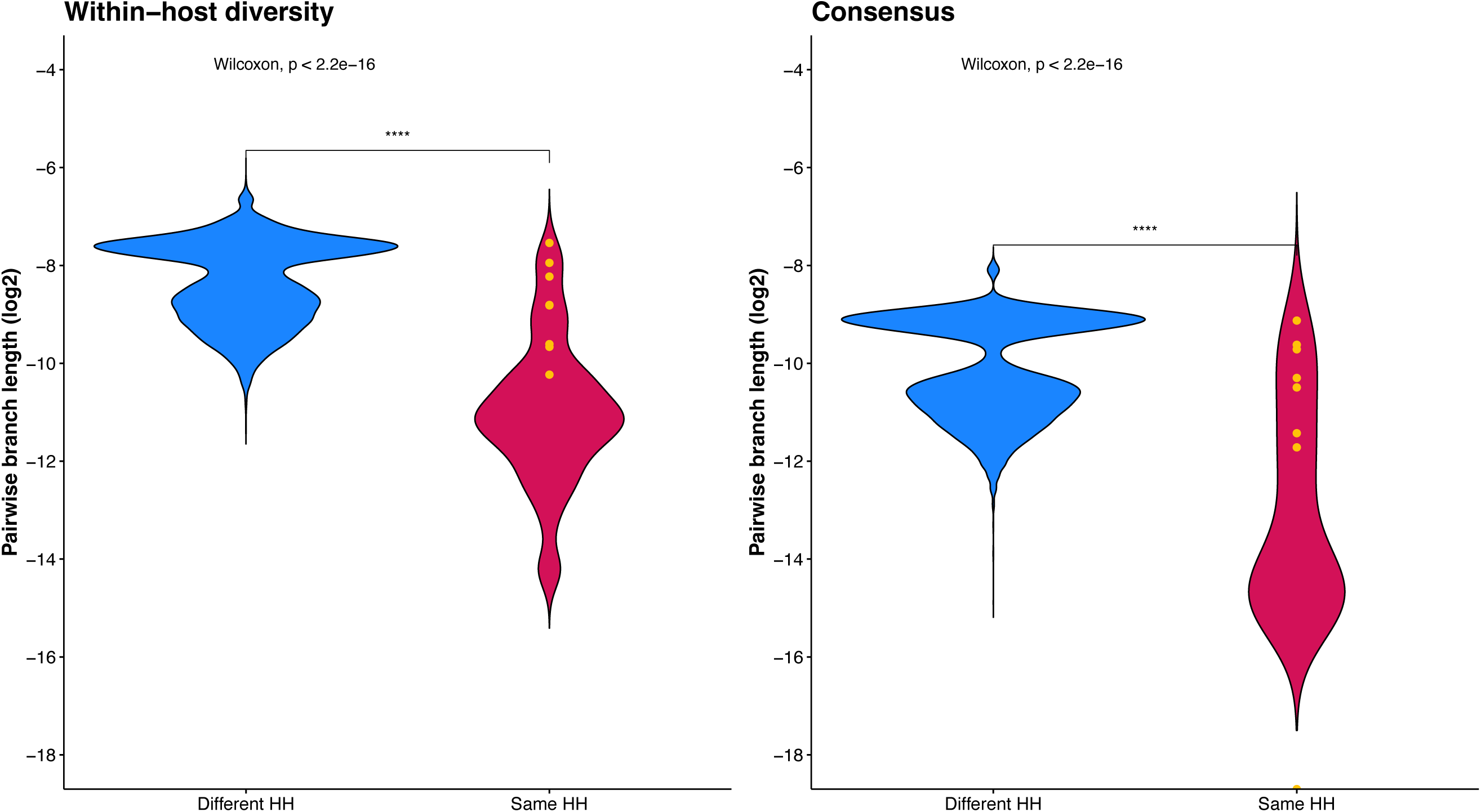
Pairwise branch distance (pairs of tips): consensus vs within-host diversity. Comparison of pairwise branch distance of SARS-CoV-2 positive samples from same/different households (HH) when considering consensus or within-host diversity phylogenetic tree. Branch length is significantly different between the two models (Wilcoxon test: p<10^-5^). Yellow dots represent households identified by phylogeny to potentially be introduction events.

**Figure 3.**
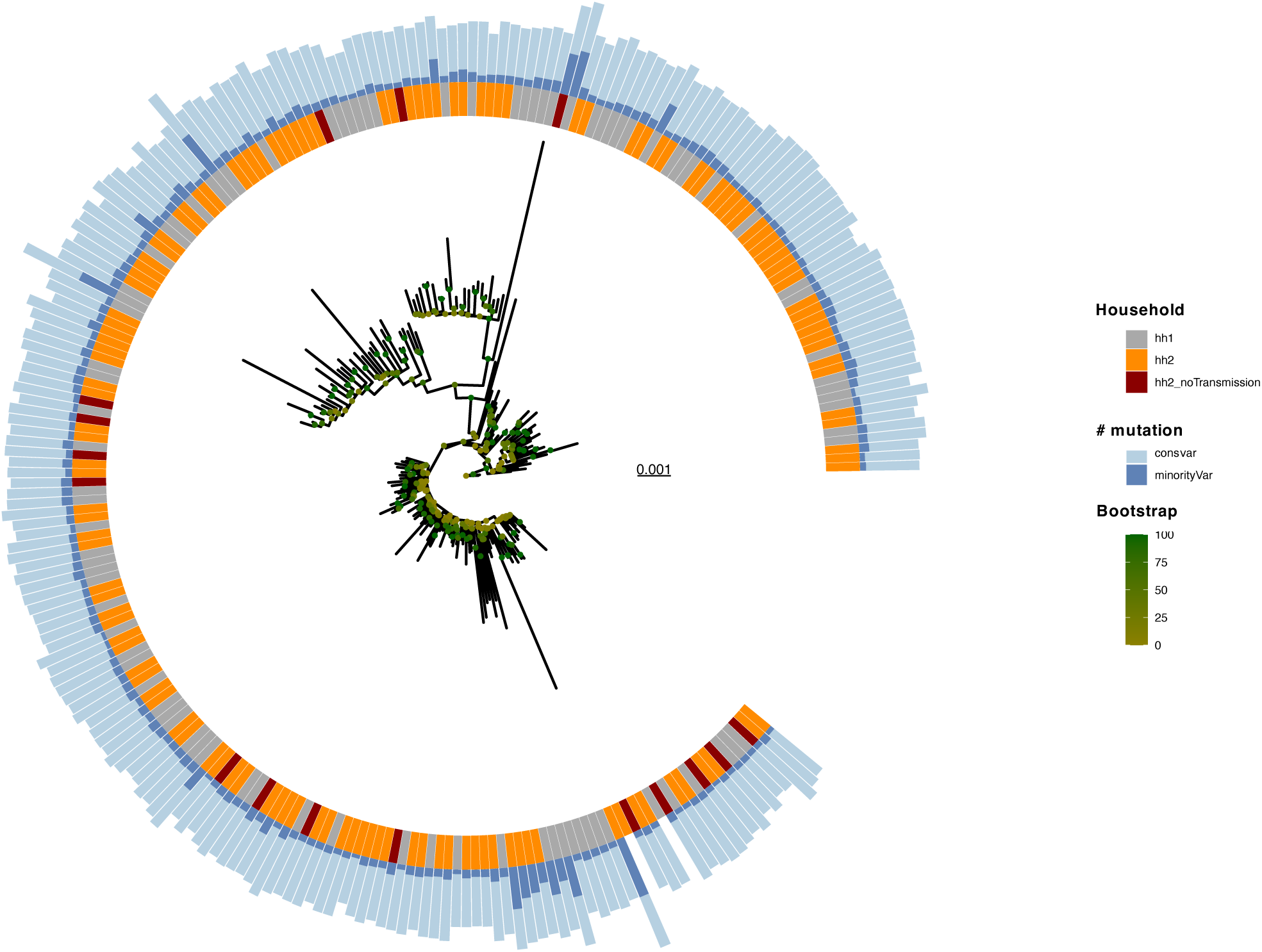
Maximum likelihood tree built by using the within-host diversity. Scale and branch lengths represent genetic distance. The inner bars show the number of positive SARS-CoV-2 cases per household. The outer bars indicate the number of consensus and low frequency variants. Branches are colored by the estimated bootstrap value.

### Secondary attack rate

Excluding the eight contacts where household transmission had been ruled out we calculated a secondary attack rate (SAR) of between 0.2 and 0.5 (mean 0.45, 95% CI [0.42,0.47]). To further define index/donor cases when the collection date of the samples from the same household was identical (41.5% of the households with more than one positive participant) we used the within-host maximum likelihood tree, to generate a dated phylogenetic tree^26^ as well as consider both the proportion of variants shared between the index and recipient sequences in each transmission pair and the posterior probability of transmission (Supplementary Figure 6). Where the index case collection date clearly preceded that of the recipient, we found the latter to have a higher proportion of shared variants in 76% of the cases (p-value = 0.0229 from 10,000 permutation tests) as would be expected^11,32^ following a transmission bottleneck. We therefore used the proportion of shared variants to assign the index and recipient status in cases where the collection date of two or more positive samples was the same (Figure 4b). The transmission window interval, i.e. the number of days between index and recipient testing positive, was between 1 to 9 days with only one outlier of 20 days. For the eight households where phylogenetic analysis excluded household transmission, the apparent transmission window interval was wider spanning from 1 to 130 days. There were 153 primary cases and 200 other members of households to which the primary cases belonged (Tables 2 and 3 present their baseline characteristics). The relative risk of a primary case transmitting the infection to one or more other household members was 0.98 with each successive vaccine dose (p=0.916, 95% CI=0.65-1.47). The relative risk of a household member acquiring the infection from the primary case was 0.88 with each successive vaccine dose (p=0.476, 95% CI=0.61-1.26). While the relative risks for both scenarios point in the expected direction of vaccine being protective, the evidence for this is weak due to the small sample size.

**Figure 4.**
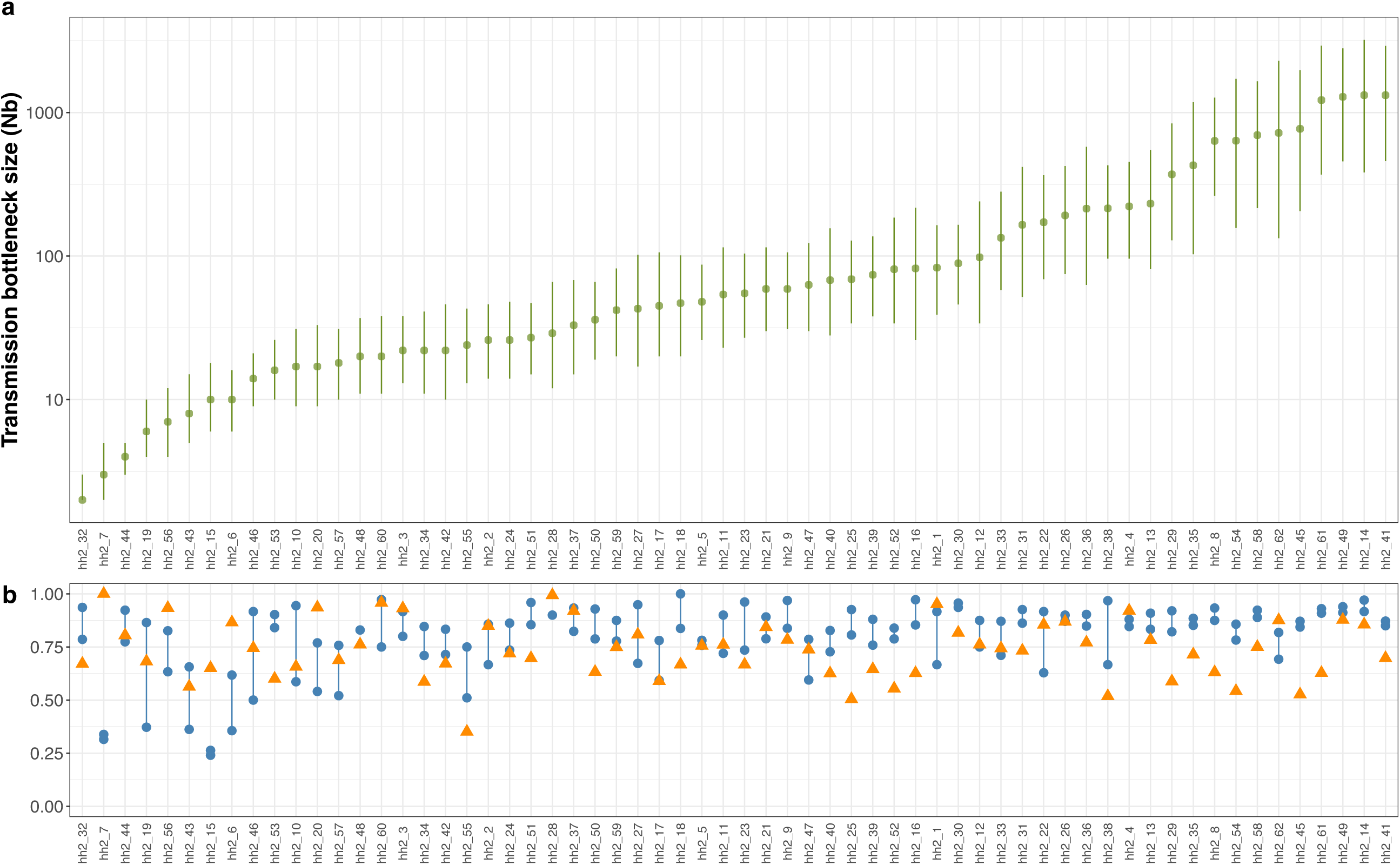
**a)** Estimated bottleneck size (as population size, *Nb*) with Koelle approximate binomial model (left Y-axis, Log10) for different households. **b)** Posterior probability estimated with TransPhylo (orange triangles) and proportion of shared variants (blue dots) per household.

**Table 2.**
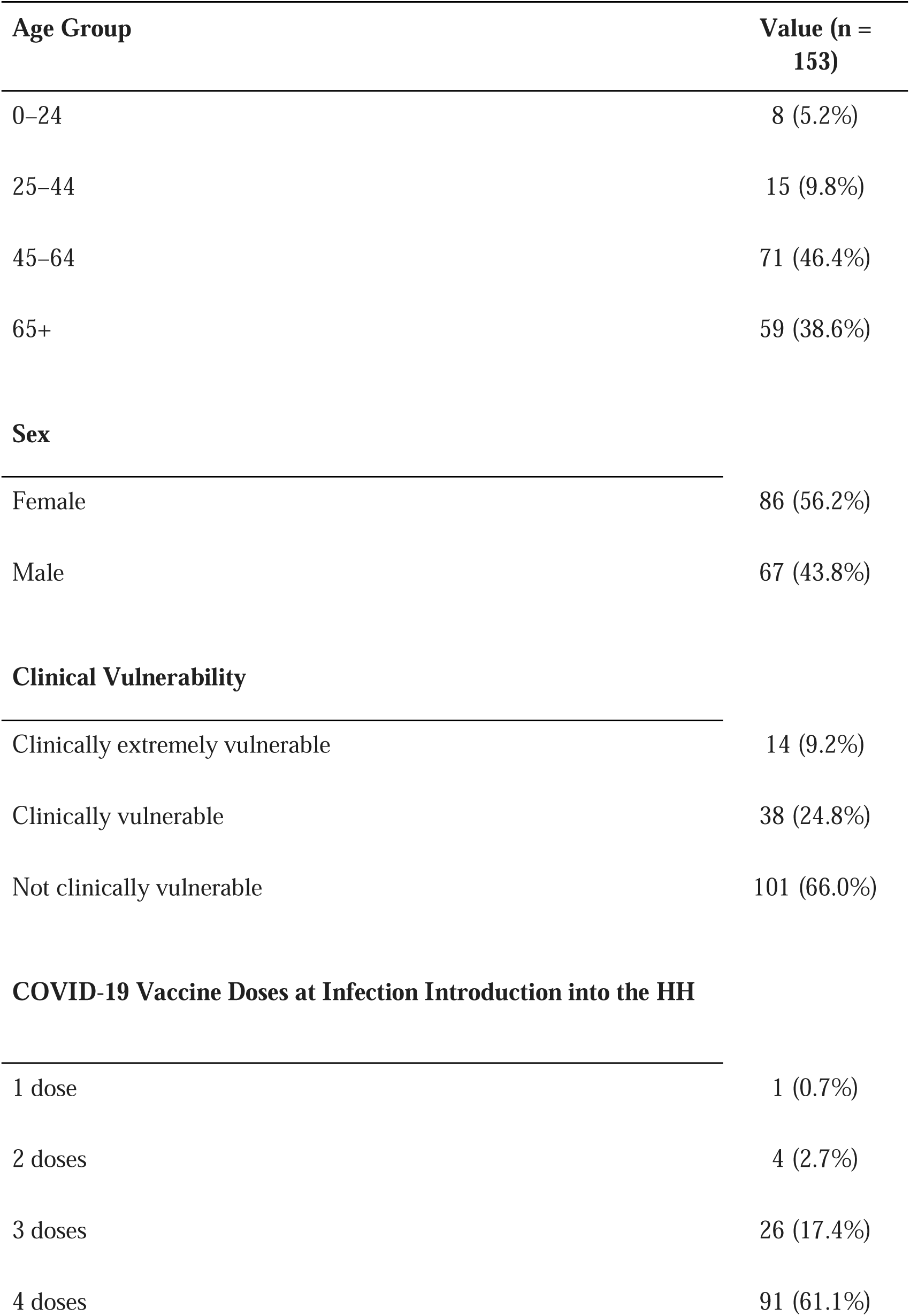
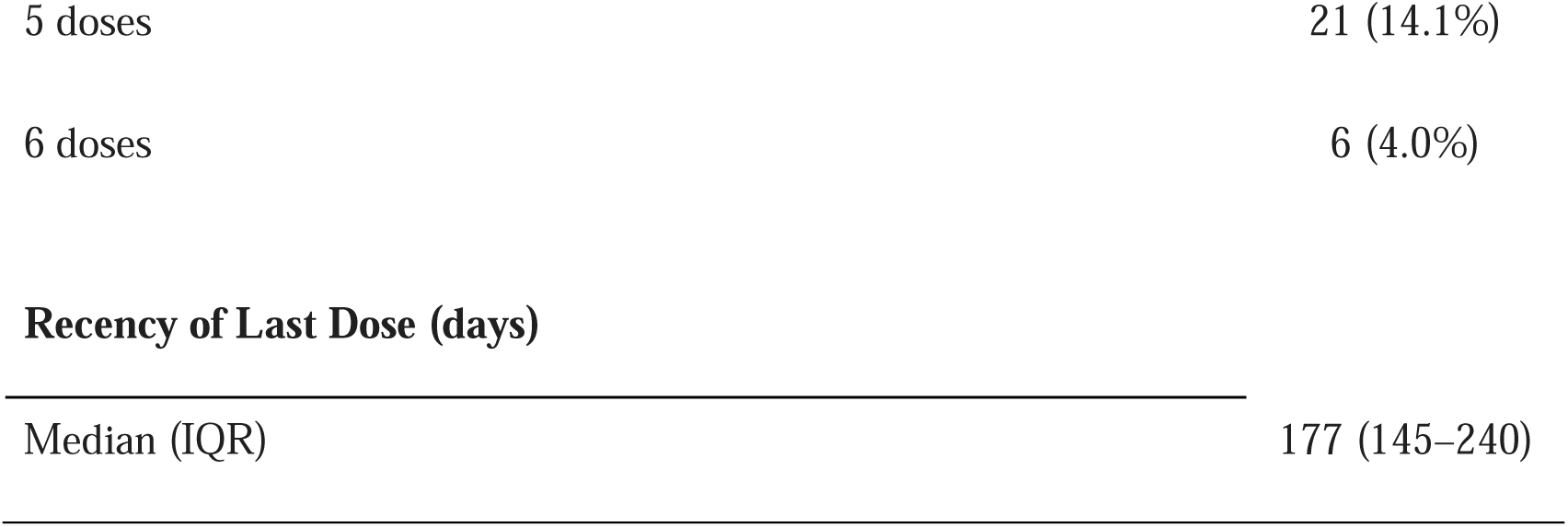
Baseline characteristics of the primary cases of households (HH).

| **Age Group** | **Value (n = 153)** |
| --- | --- |
| 0–24 | 8 (5.2%) |
| 25–44 | 15 (9.8%) |
| 45–64 | 71 (46.4%) |
| 65+ | 59 (38.6%) |
| **Sex** |  |
| Female | 86 (56.2%) |
| Male | 67 (43.8%) |
| **Clinical Vulnerability** |  |
| Clinically extremely vulnerable | 14 (9.2%) |
| Clinically vulnerable | 38 (24.8%) |
| Not clinically vulnerable | 101 (66.0%) |
| **COVID-19 Vaccine Doses at Infection Introduction into the HH** |  |
| 1 dose | 1 (0.7%) |
| 2 doses | 4 (2.7%) |
| 3 doses | 26 (17.4%) |
| 4 doses | 91 (61.1%) |
| 5 doses | 21 (14.1%) |
| 6 doses | 6 (4.0%) |
| **Recency of Last Dose (days)** |  |
| Median (IQR) | 177 (145–240) |

**Table 3.**
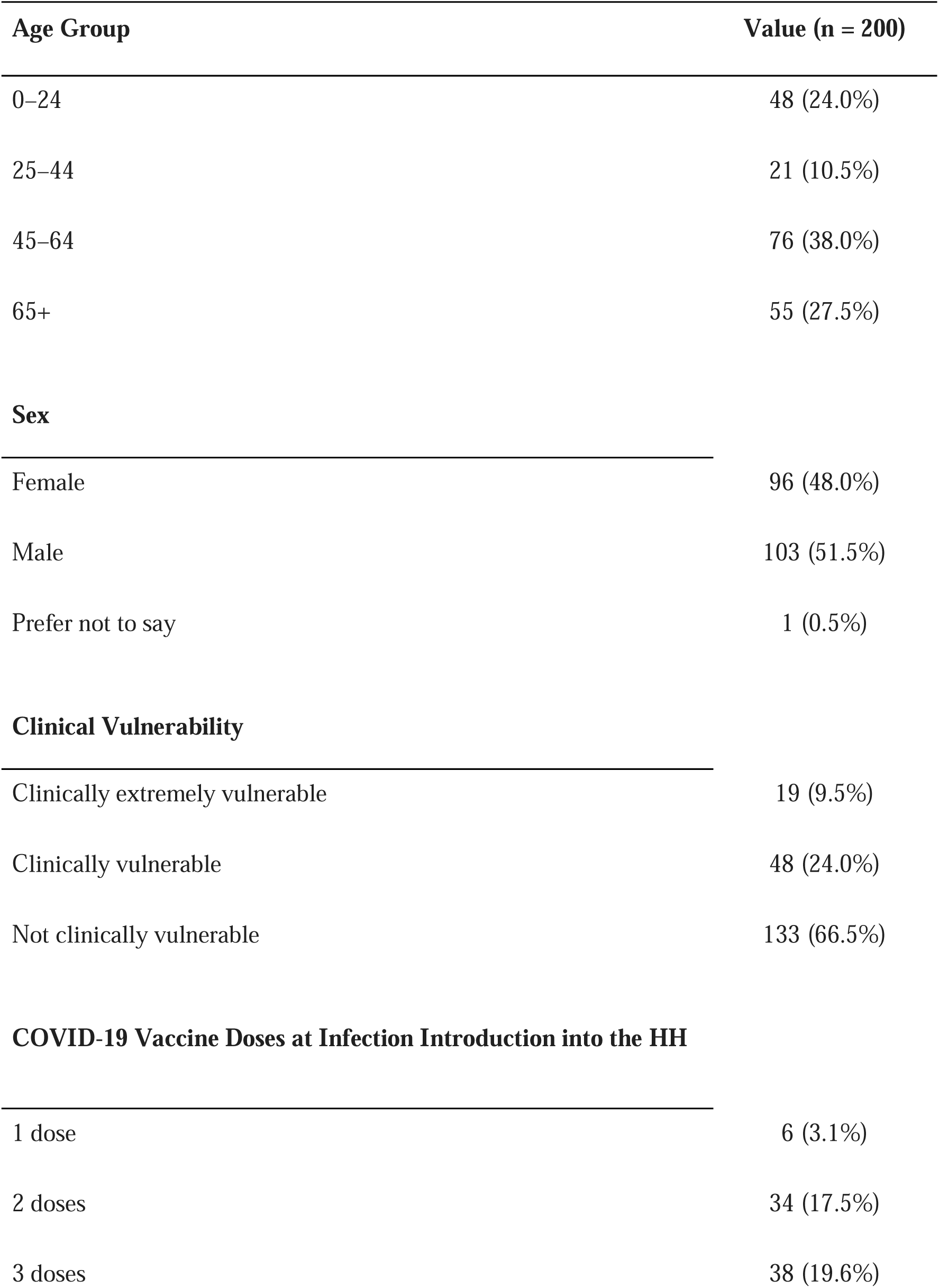
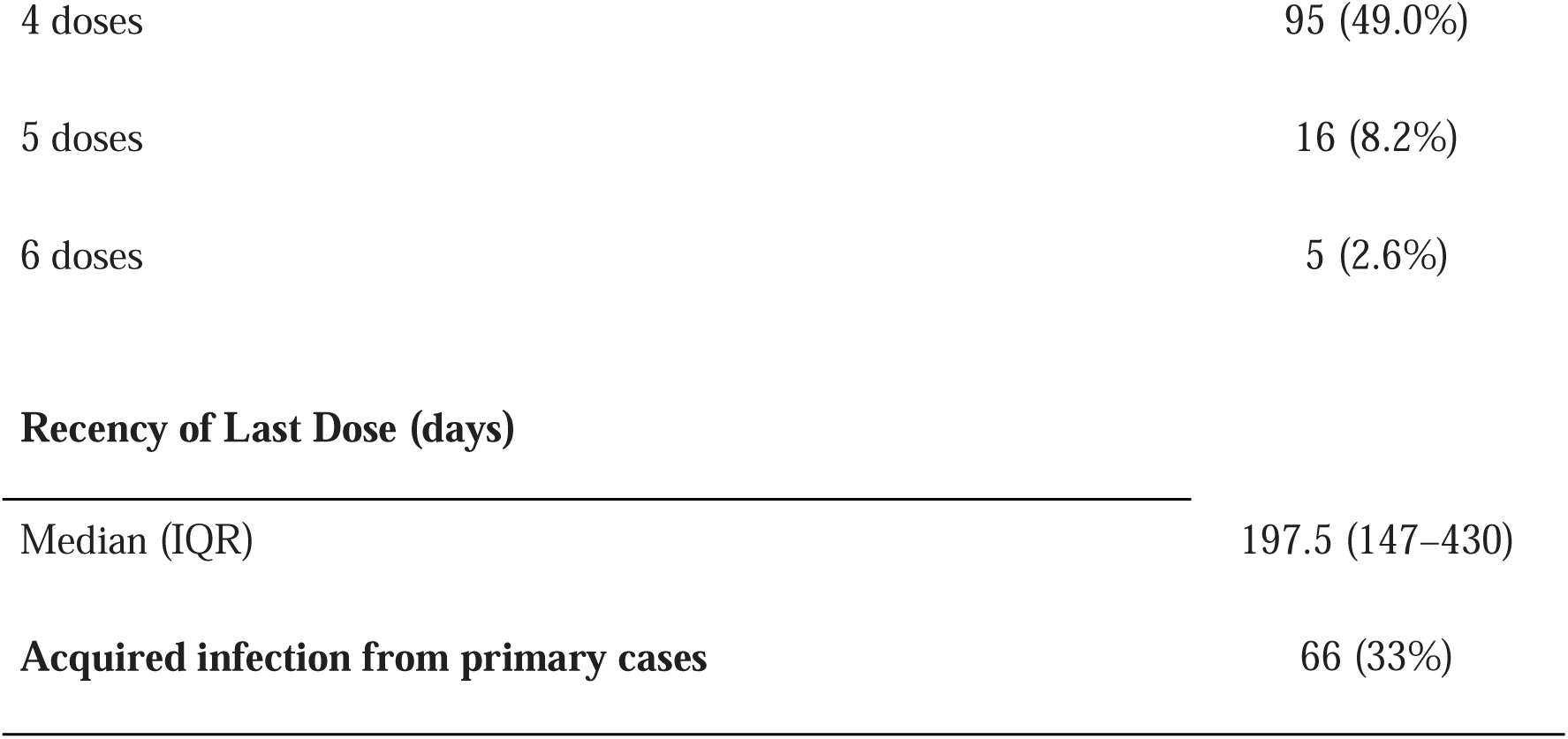
Baseline characteristics of other members of households (HH) of primary cases (potential secondary cases).

| Age Group | Value (n = 200) |
| --- | --- |
| 0–24 | 48 (24.0%) |
| 25–44 | 21 (10.5%) |
| 45–64 | 76 (38.0%) |
| 65+ | 55 (27.5%) |
| **Sex** |  |
| Female | 96 (48.0%) |
| Male | 103 (51.5%) |
| Prefer not to say | 1 (0.5%) |
| **Clinical Vulnerability** |  |
| Clinically extremely vulnerable | 19 (9.5%) |
| Clinically vulnerable | 48 (24.0%) |
| Not clinically vulnerable | 133 (66.5%) |
| **COVID-19 Vaccine Doses at Infection Introduction into the HH** |  |
| 1 dose | 6 (3.1%) |
| 2 doses | 34 (17.5%) |
| 3 doses | 38 (19.6%) |
| 4 doses | 95 (49.0%) |
| 5 doses | 16 (8.2%) |
| 6 doses | 5 (2.6%) |
| **Recency of Last Dose (days)** |  |
| Median (IQR) | 197.5 (147–430) |
| **Acquired infection from primary cases** | 66 (33%) |

### Bottleneck size estimation

The bottleneck size was estimated using the approximate beta-binomial method^34^, incorporating within-host diversity. The estimated overall bottleneck size had a mean of 202 (95% CI: [115, 290]) virions. Within each household, possible transmission pairs included the index case as the donor and each confirmed household contact as a recipient (the eight household contacts with unrelated viruses were excluded; Figure 4a). Moreover, we compared the number of shared variants (allele frequency intervals: 5-95% and 10-95%) between inferred transmission pairs and random pairs and in both scenarios random pairs had significantly lower number of shared mutations (Supplementary Figure 7).

## Discussion

Whole genome sequencing provides a powerful tool for identifying related infections. However, where mutation rates are low, as for SARS-CoV-2, the lack of sequence variation in the timescale during which household transmissions are occurring can limit its utility as previously described^5,7,8,35^. Using novel phylogenetic methods that make use of minority variants^11^, we were able more accurately to distinguish household from community transmission than could be achieved if only consensus sequences had been used. While consensus phylogeny identified clustering of cases within the same household, transmission directionality was resolved only when using within-host diversity. Analysis of over 4,000 consensus GISAID sequences suggested sufficient SARS-CoV-2 diversity to exclude spurious household clustering due to a recent selective sweep, a finding in keeping with evidence that all the Omicron variants represented in this study had been circulating for at least three months (https://covid19.sanger.ac.uk). This observation may also reflect the fact that COVID-19 testing was no longer mandatory at the time, and that the available sequence data may not fully represent the UK communities surrounding the Virus Watch households. While in the majority of households with more than one positive participant, cases clustered together, we were able to exclude eight putative household inferred transmission events. Among the four pairs with clear differences in the Omicron lineage between index and recipient, three had a wide window of infection, between 14 and 38 days, further supporting a likely different source of infection.

The dated phylogeny generated using the within-host diversity model together with the higher proportion of shared variants agreed with the samples’ collection dates in 76% of the cases where the collection date was different for the household’s members (p-value: 0.023). Of note, the sample collection date is not expected to be a perfect indicator of transmission directionality, with a recent study^36^ having found a negative serial interval in about 20% of transmission events. From this dataset we calculate the household attack rate to be slightly lower than has previously been observed including for Omicron variants, but consistent with the results of other studies^37–40^.

In our multivariate analyses, we observed only weak evidence suggesting that an increased number of vaccine doses may be associated with a reduced risk of acquiring infection from the household index case. However, this association was not statistically robust and should be interpreted with caution. The limited sample size in our study likely reduced the power to detect modest effects. While some previous studies have reported a protective effect of vaccination against secondary transmission within households^41,42^, our findings highlight the challenges in assessing this relationship in smaller cohorts.

In contrast to other SARS-CoV-2 household transmission studies^2,3,43^, our approach combined epidemiological data with genomic sequencing, including within-host diversity, to improve identification of index cases and infer transmission direction. Nonetheless, our study has limitations. Index cases were identified following a positive test and self-reported symptoms, which may underestimate the secondary infection rate if earlier, asymptomatic infections went undetected. Furthermore, the time lag between the index case’s positive test and enrolment of other household members likely limited our ability to precisely capture the timing of secondary infections, as only symptomatic individuals were prompted to perform a lateral flow test, participants testing positive shortly after enrolment may have acquired infection prior to the assumed index case. These constraints could be addressed by prospective household enrolment prior to infection and systematic, symptom independent testing; however, such a design poses significant logistical challenges.

An interesting corollary to these data is our understanding of the SARS-CoV-2 transmission bottleneck. Previous studies using a similar approach but considering only the consensus data have estimated a tight bottleneck for SARS-CoV-2^8,10,12,44^, by contrast our data suggest greater sharing of unique minority variants between sequences from household contacts when within-host diversity was also considered. We also observed a significantly lower fraction of minority variants shared between unrelated samples (p<0.0074). Taken together, viral genome sequencing and analyses that incorporate minority variant data, can improve the identification of within household transmissions events, and suggest that infection may be established from the transmission of anywhere between 2 and 1,321 virions, reflecting a wide range of transmission bottleneck sizes.

## Materials and Methods

### Sample collection and processing

Participants in the genomic sub-cohort were recruited through a randomized sample of individuals who had previously participated in the Vaccine Watch study^18^, along with their household members, resulting in a total of 3,000 participants. Lead householders received an invitation email and were asked to review the participant information sheets and discuss the study with their household. Informed consent was obtained through an online form completed by each adult, while children provided assent with parental or guardian support. During the six-month follow-up period (January 1 – June 30, 2023), participants were instructed to perform a lateral flow test if they experienced fever, new continuous cough, or a loss or altered sense of smell or taste. If a test was positive, participants were asked to self-collect a swab sample and return it via reply-paid mail to the Camelia Botnar Laboratories at Great Ormond Street Hospital for Children NHS Foundation Trust for further analysis.

Upon arrival at the Great Ormond Street Hospital laboratory, the swabs in VTM were agitated on a vortex mixer, and 250 µl of the sample fluid was transferred to a sterile Sarstedt tube containing lysis buffer for RNA extraction using the Microlab STAR platform. To control for extraction failures, PDV (Phocine Distemper Virus) was spiked into every sample during the extraction process. Negative sample extraction controls were included in each batch of extractions. After extraction, the samples were tested for SARS-CoV-2 and PDV using multiplex targeted one-step real-time RT-PCR on a Quantstudio 5 instrument. Each PCR run included a SARS-CoV-2 positive template control and a no-template control. Extraction and PCR control results were recorded and monitored for consistency.

### Viral Load

Viral loads were determined using a standard curve generated from a ten-fold dilution series of SARS-CoV-2 RNA with a known quantity. This curve was imported into the PCR analysis to calculate the viral load in copies/mL for each positive sample. The lower limit of quantification (LLOQ) was set at 109 copies/mL corresponding to a Ct value of 40. RNA samples positive for SARS-CoV-2 with a Ct value below 38 were subsequently sent to UCL Genomics for sequencing.

### Sequencing

Libraries were prepared on the Agilent Bravo liquid handling robot using the CoronaHiT^19^ protocol for Illumina sequencing and ARTIC v4.1 primers. Sequencing was performed with a target depth of 5000X per genome on an Illumina sequencer using 2x150-bp paired-end reads. All runs included both positive and negative controls to detect contamination at each step of the protocol.

### Sequence Analysis

The raw fastq reads were adapted, trimmed and low-quality reads removed using the fastp algorithm. The reads were aligned against the Wuhan-Hu-1 reference genome (NC_045512.2, MN908947) sequence and then the amplicon primer regions were trimmed using the location provided in a bed file. Consensus sequences were called at a minimum of 30× coverage. The entire processing of raw reads to consensus was carried out using nf-core/viralrecon pipeline^20^ (https://nf-co.re/viralrecon/2.6.0), where Pangolin^21^ was used to assign SARS-CoV-2 lineages. Only samples producing genomes with at least 90% genome coverage at 10× sequencing depth were kept for further analysis. Variant calling was done using the iVAR algorithm^22^. Variants were further filtered using bcftools^23^. Only variants with a minimum depth of 50× and a minimum base quality and mapping quality of 30 were retained.

Previously identified problematic sites were masked to avoid systematic sequencing errors and phylogenetic bias (https://github.com/W-L/ProblematicSites_SARS-CoV2). Moreover, variants present in more than 80% of the population were also masked. At positions with no minority variants, the minimum depth was 20 reads, with at least 5 reads in each direction. Positions with low-frequency variants were filtered if the total coverage at that position was less than 200×, with at least 20 reads in total and at least 5 independent reads with no strand-bias supporting each of the main two alleles.

### Phylogenetic analysis

Two different alignments were prepared from the data following Torres Ortiz *et al*. 2023^11^: 1) a consensus sequence alignment where only the majority allele is represented; and 2) an alignment that keeps low frequency variants with a minimum allele frequency of 1%. For the two different alignments, maximum likelihood phylogenies were inferred using RAxML-NG^24^ with 20 starting trees (10 random and 10 parsimony), 1000 bootstrap replicates, and a minimum branch length of 10^-9^. The phylogeny with the consensus alignment was inferred using a GTR+G model. A model with 16 states was used to infer the tree from the low frequency variant alignment, where all states were allowed to have their own rate^11^.

Phylogenetic trees were time-calibrated using the known collection dates and the additive relaxed clock (ARC) model^25^ within BactDating^26^. For transmission inference, the dated phylogeny was used as input. Posterior probabilities of direct transmission were estimated using TransPhylo^27^ for one million MCMC iterations. The generation time was a Gamma distribution with mean and standard deviation 0.01 year and 0.009 year respectively. Having the standard deviation slightly lower than the mean implies that the shape of the Gamma distribution is greater than one, so that infectivity starts at zero before increasing up to a peak and then decreasing. The mean corresponds to 3.65 days, with the 95% quantile being at 12 days, which is compatible with previous estimates of SARS-CoV-2 generation time^28^. We also applied TransPairs^29^ to compute the likelihood of each potential directed transmission event, with a mean latency period of 4 days and mean infectious period of 6 days. A total of 4219 high coverage consensus sequences generated between January to June 2023 were downloaded from Nextstrain^30^ and used for comparative analysis. General data processing was carried out in R4.2.0, figures were made using the ggplot2 package^31^.

### Secondary attack rate

Among households with at least one positive member, the index case was defined as the earliest confirmed positive in the household (based on the sample collection date), secondary cases as household contacts who subsequently became positive with SARS-CoV-2 genome sequences clustered phylogenetically with the index, and negatives as those household contacts who either did not experience any of the symptoms described in the methods or the lateral flow test was negative throughout the duration of the study. In cases where the collection date for participants from the same household was the same and the genome sequences clustered together, we used both the dated phylogeny and the proportion of shared variants to infer index and secondary case, assuming that the recipient has a higher proportion of shared variants^11,32^. Secondary attack rate (SAR) was calculated as the proportion of total number of secondary infections among all susceptible contacts per household.

We modelled two transmission scenarios: the risk of a household introduction (primary) case infecting one or more other household members (secondary cases), and the risk of a negative household member acquiring the infection from the index, with the number of vaccination doses being the exposure. For both models, we used mixed-effects poisson regression for binary outcomes with a random intercept at the household level to account for clustering within households. Additionally, we used robust errors to improve model parameter estimation^33^. We modelled the number of doses as an ordinal variable, where the minimum number of doses is 1 (we did not consider unvaccinated individuals). The covariates for both models were the recency of the last dose (in days), age, sex and clinical vulnerability status.

### Bottleneck estimation

We defined the possible transmission pairs within each household using the criteria for secondary attack rate (SAR) outlined above. After defining the transmission pairs, we applied the approximate beta-binomial (ABB) model^34^. The ABB model extends the standard binomial distribution by incorporating a beta-distributed prior on the binomial probability parameter, effectively capturing extra-binomial variation. This approach allows for greater flexibility in modelling biological and epidemiological data where variance exceeds the mean, which is commonly observed in viral evolution and immune escape dynamics. We estimated the bottleneck size for each transmission pair using the following variant calling thresholds: 1%, 3% and 5% (Supplementary Figure 1).

## Data availability

The raw data used in this study have been deposited in the Office of National Statistics (ONS) Secure Research Service. Access to the data is restricted to the five safe’s framework consisting of: safe data, safe projects, safe people, safe settings and safe outputs. The institution review board which assesses appropriateness of the researcher, and the project is the Office of National Statistics and as part of the data application process, researchers are expected to participate in the ONS “People and Projects Service” and project accreditation.

Access to the Virus Watch data can be found: https://doi.org/10.57906/s5f5-nq13

## Code availability

Associated code is publicly available on GitHub ().

## Acknowledgements

We thank all individuals who participated in this study and Sarah Beale for useful comments. This work was supported by the Medical Research Council [Grant Ref: MC_PC 19070] awarded to UCL on 30 March 2020 and Medical Research Council [Grant Ref: MR/V028375/1] awarded on 17 August 2020. The study also received $15,000 of advertising credit from Facebook to support a pilot social media recruitment campaign on 18th August 2020.

This study was supported by the Wellcome Trust through a Wellcome Clinical Research Career Development Fellowship to R.W.A. [206602].

From 1 May 2022 Virus Watch received funding from the European Union (Project: 101046314). Views and opinions expressed are however those of the author(s) only and do not necessarily reflect those of the European Union or the European Health and Digital Executive Agency (HaDEA). Neither the European Union nor the granting authority can be held responsible for them.

## Contributions

J.B. and L.B. contributed to study concept and design; L.B. and A.T.O. performed phylogenetic and bioinformatics analyses; J.B.,A.T.O.,X.D., and L.B. contributed to data interpretation; L.M.M.B. performed the genomic sequencing; R.W. provided management and oversight of the laboratory staff; C.M. contributed to sample collection and sample testing; J.K., A.Y., and C.G. provided management and oversight for the cohort project; A.Y and L.B. contributed to data curation; A.Y. performed epidemiologic analyses; supervision, J.B; funding acquisition, R.W.A and I.A.; L.B. and J.B. wrote the original draft, with all authors reviewing and editing.

## Reference

1. Cerami, C. et al. Household Transmission of Severe Acute Respiratory Syndrome Coronavirus 2 in the United States: Living Density, Viral Load, and Disproportionate Impact on Communities of Color. Clin. Infect. Dis. Off. Publ. Infect. Dis. Soc. Am. 74, 1776–1785 (2022).

2. Hare, D. et al. Genomic epidemiological analysis of SARS-CoV-2 household transmission. Access Microbiol. 3, 000252 (2021).

3. Kolodziej, L. M. et al. High Severe Acute Respiratory Syndrome Coronavirus 2 (SARS-CoV-2) Household Transmission Rates Detected by Dense Saliva Sampling. Clin. Infect. Dis. Off. Publ. Infect. Dis. Soc. Am. 75, e10 (2022).

4. Agoti, C. N. et al. Transmission networks of SARS-CoV-2 in Coastal Kenya during the first two waves: A retrospective genomic study. eLife 11, e71703 (2022).

5. Jelley, L. et al. Tracing household transmission of SARS-CoV-2 in New Zealand using genomics. Npj Viruses 2, 1–8 (2024).

6. Bendall, E. E., et al. SARS-CoV-2 Genomic Diversity in Households Highlights the Challenges of Sequence-Based Transmission Inference. mSphere 7, e0040022 (2022).

7. Lyngse, F. P. et al. Household transmission of SARS-CoV-2 Omicron variant of concern subvariants BA.1 and BA.2 in Denmark. Nat. Commun. 13, 5760 (2022).

8. Bendall, E. E. et al. Rapid transmission and tight bottlenecks constrain the evolution of highly transmissible SARS-CoV-2 variants. Nat. Commun. 14, 272 (2023).

9. Biek, R., Pybus, O. G., Lloyd-Smith, J. O. & Didelot, X. Measurably evolving pathogens in the genomic era. Trends Ecol. Evol. 30, 306–313 (2015).

10. Tonkin-Hill, G. et al. Patterns of within-host genetic diversity in SARS-CoV-2. eLife 10, e66857 (2021).

11. Torres Ortiz, A., et al. Within-host diversity improves phylogenetic and transmission reconstruction of SARS-CoV-2 outbreaks. eLife 12, e84384 (2023).

12. Lythgoe, K. A. et al. SARS-CoV-2 within-host diversity and transmission. Science 372, eabg0821 (2021).

13. Markov, P. V. et al. The evolution of SARS-CoV-2. Nat. Rev. Microbiol. 21, 361–379 (2023).

14. Braun, K. M. et al. Acute SARS-CoV-2 infections harbor limited within-host diversity and transmit via tight transmission bottlenecks. PLOS Pathog. 17, e1009849 (2021).

15. Liu, Y., Eggo, R. M. & Kucharski, A. J. Secondary attack rate and superspreading events for SARS-CoV-2. Lancet Lond. Engl. 395, e47 (2020).

16. Lemieux, J. E. et al. Phylogenetic analysis of SARS-CoV-2 in Boston highlights the impact of superspreading events. Science 371, eabe3261 (2021).

17. Byrne, T. et al. Cohort Profile: Virus Watch-understanding community incidence, symptom profiles and transmission of COVID-19 in relation to population movement and behaviour. Int. J. Epidemiol. 52, e263–e272 (2023).

18. Byrne, T. et al. Trends, patterns and psychological influences on COVID-19 vaccination intention: Findings from a large prospective community cohort study in England and Wales (Virus Watch). Vaccine 39, 7108–7116 (2021).

19. Baker, D. J. et al. CoronaHiT: high-throughput sequencing of SARS-CoV-2 genomes. Genome Med. 13, 21 (2021).

20. Patel, H. et al. nf-core/viralrecon: nf-core/viralrecon v2.6.0 - Rhodium Raccoon. (2023).

21. Rambaut, A. et al. A dynamic nomenclature proposal for SARS-CoV-2 lineages to assist genomic epidemiology. Nat. Microbiol. 5, 1403–1407 (2020).

22. Grubaugh, N. D. et al. An amplicon-based sequencing framework for accurately measuring intrahost virus diversity using PrimalSeq and iVar. Genome Biol. 20, 8 (2019).

23. Danecek, P. et al. The variant call format and VCFtools. Bioinforma. Oxf. Engl. 27, 2156–2158 (2011).

24. Kozlov, A. M., Darriba, D., Flouri, T., Morel, B. & Stamatakis, A. RAxML-NG: a fast, scalable and user-friendly tool for maximum likelihood phylogenetic inference. Bioinforma. Oxf. Engl. 35, 4453–4455 (2019).

25. Didelot, X., Siveroni, I. & Volz, E. M. Additive Uncorrelated Relaxed Clock Models for the Dating of Genomic Epidemiology Phylogenies. Mol. Biol. Evol. 38, 307–317 (2021).

26. Didelot, X., Croucher, N. J., Bentley, S. D., Harris, S. R. & Wilson, D. J. Bayesian inference of ancestral dates on bacterial phylogenetic trees. Nucleic Acids Res. 46, e134 (2018).

27. Didelot, X., Kendall, M., Xu, Y., White, P. J. & McCarthy, N. Genomic Epidemiology Analysis of Infectious Disease Outbreaks Using TransPhylo. Curr. Protoc. 1, e60 (2021).

28. Knight, J. & Mishra, S. Estimating effective reproduction number using generation time versus serial interval, with application to covid-19 in the Greater Toronto Area, Canada. Infect. Dis. Model. 5, 889–896 (2020).

29. Eldholm, V. et al. Impact of HIV co-infection on the evolution and transmission of multidrug-resistant tuberculosis. eLife 5, e16644 (2016).

30. Hadfield, J. et al. Nextstrain: real-time tracking of pathogen evolution. Bioinforma. Oxf. Engl. 34, 4121–4123 (2018).

31. Wickham, H. Ggplot2: Elegant Graphics for Data Analysis. (Springer international publishing, Cham, 2016).

32. Kundu, S. et al. Next-generation whole genome sequencing identifies the direction of norovirus transmission in linked patients. Clin. Infect. Dis. Off. Publ. Infect. Dis. Soc. Am. 57, 407–414 (2013).

33. Zou, G. A modified poisson regression approach to prospective studies with binary data. Am. J. Epidemiol. 159, 702–706 (2004).

34. Sobel Leonard, A., Weissman, D. B., Greenbaum, B., Ghedin, E. & Koelle, K. Transmission Bottleneck Size Estimation from Pathogen Deep-Sequencing Data, with an Application to Human Influenza A Virus. J. Virol. 91, e00171–17 (2017).

35. Allen, H. et al. Household transmission of COVID-19 cases associated with SARS-CoV-2 delta variant (B.1.617.2): national case-control study. Lancet Reg. Health - Eur. 12, 100252 (2022).

36. Geismar, C. et al. Bayesian reconstruction of household transmissions to infer the serial interval of COVID-19 by variants of concern: analysis from a prospective community cohort study (Virus Watch). The Lancet 400, S40 (2022).

37. Ito, K., Piantham, C. & Nishiura, H. Relative instantaneous reproduction number of Omicron SARS-CoV-2 variant with respect to the Delta variant in Denmark. J. Med. Virol. 94, 2265–2268 (2022).

38. Jørgensen, S. B., Nygård, K., Kacelnik, O. & Telle, K. Secondary Attack Rates for Omicron and Delta Variants of SARS-CoV-2 in Norwegian Households. JAMA 327, 1610–1611 (2022).

39. Kang, S.-W. et al. Comparison of secondary attack rate and viable virus shedding between patients with SARSLCoVL2 Delta and Omicron variants: A prospective cohort study. J. Med. Virol. 95, e28369 (2022).

40. Jalali, N. et al. Increased household transmission and immune escape of the SARS-CoV-2 Omicron compared to Delta variants. Nat. Commun. 13, 5706 (2022).

41. Harris, R. J. et al. Effect of Vaccination on Household Transmission of SARS-CoV-2 in England. N. Engl. J. Med. 385, 759–760 (2021).

42. Madewell, Z. J., Yang, Y., Longini, I. M., Halloran, M. E. & Dean, N. E. Household Secondary Attack Rates of SARS-CoV-2 by Variant and Vaccination Status: An Updated Systematic Review and Meta-analysis. *JAMA Netw*. Open 5, e229317 (2022).

43. Banga, J. et al. Severe Acute Respiratory Syndrome Coronavirus 2 Household Transmission During the Omicron Era in Massachusetts: A Prospective, Case-Ascertained Study Using Genomic Epidemiology. Open Forum Infect. Dis. 11, ofae591 (2024).

44. Hannon, W. W. et al. Narrow transmission bottlenecks and limited within-host viral diversity during a SARS-CoV-2 outbreak on a fishing boat. Virus Evol. 8, veac052 (2022).

