## Supplementary figures for "Reconstruction of SARS-CoV-2 transmissibility within households in the UK Virus Watch Study"

**Supplementary Figure 1.** Density plot of the estimated bottleneck size (as population size, $N_b$) with Koelle approximate binomial model (X-axis, Log10) using 1% (light blue), 3% (light pink) and 5% (light green) thresholds.

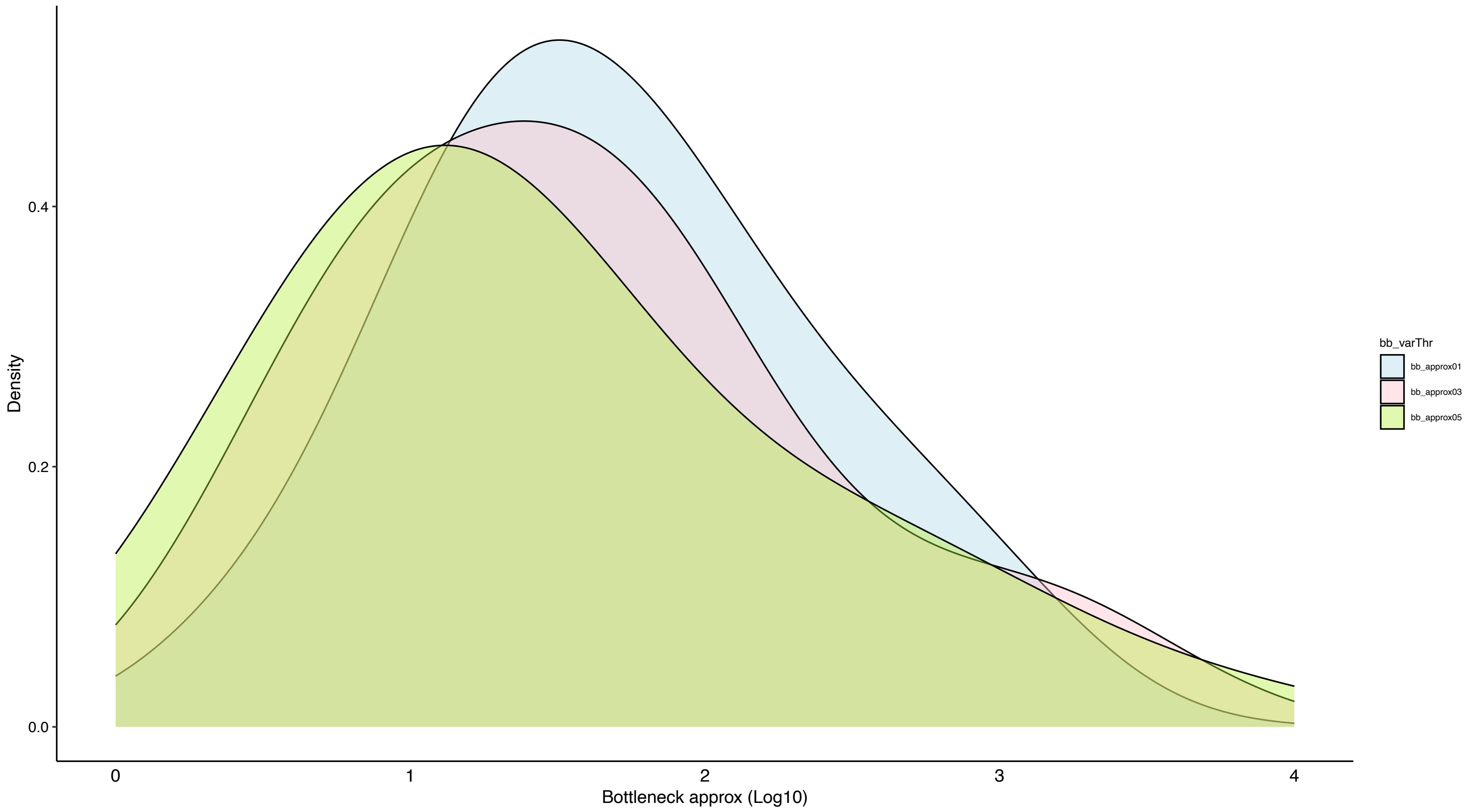

**Supplementary Figure 2.** Household distribution in the UK. Different colors represent various districts/regions. Dots represent households with one positive participant while triangles represent households with more than one positive participant.

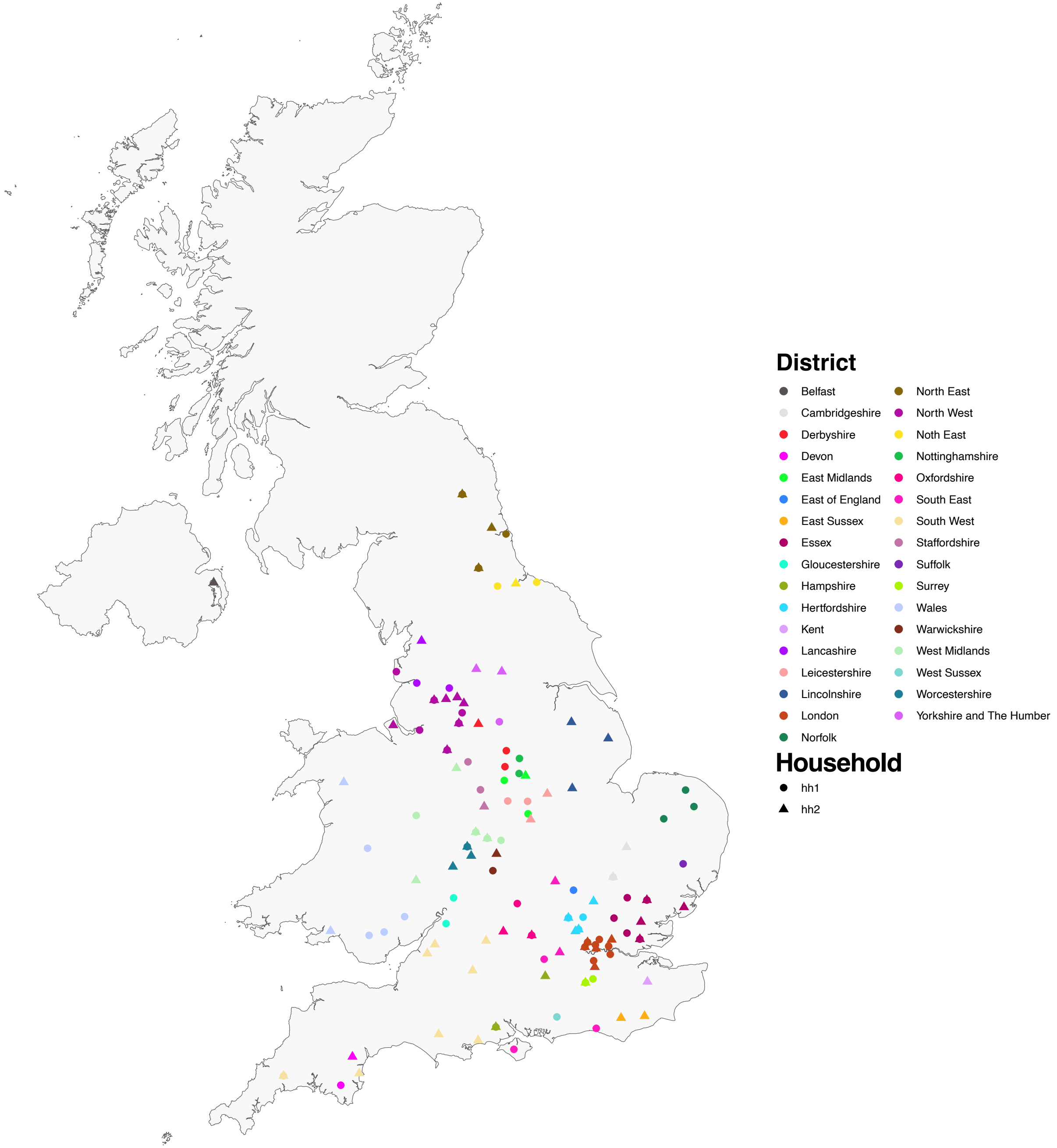

**Supplementary Figure 3.** Lineages distribution in UK *vs* genomic Virus Watch cohort.

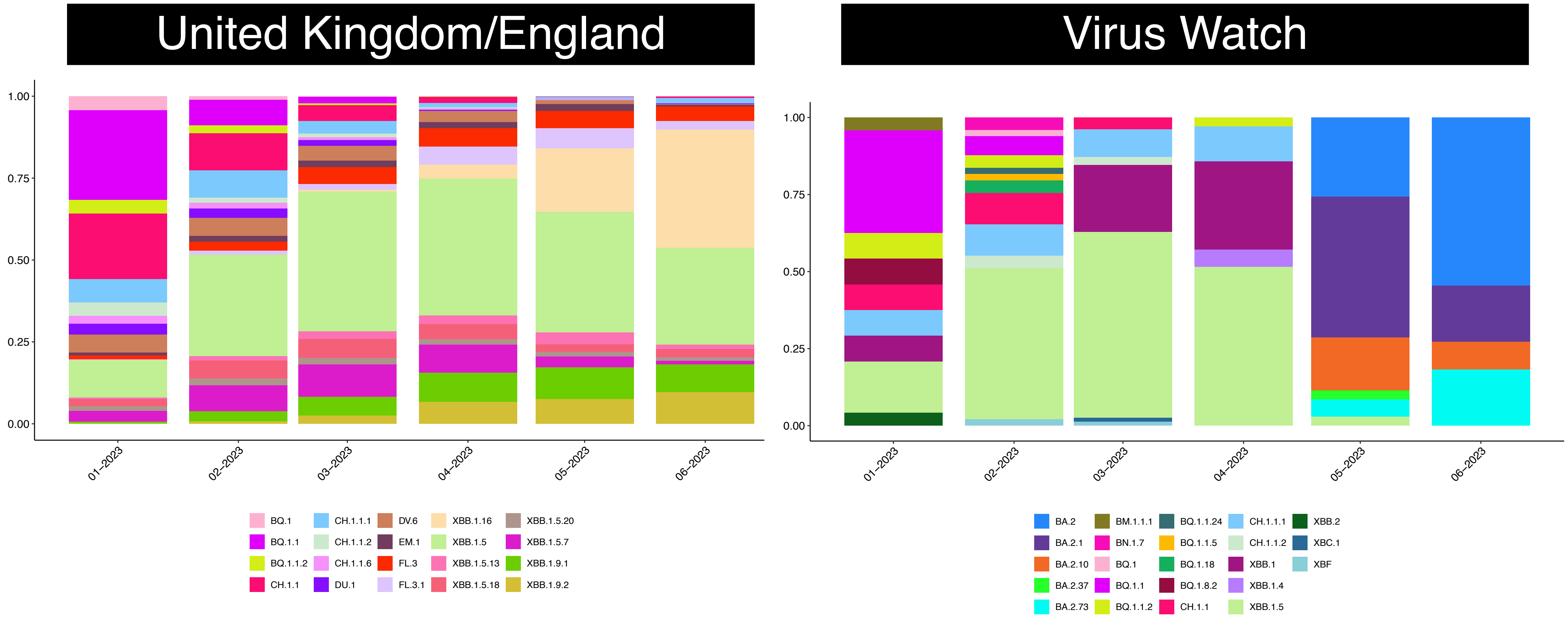

**Supplementary Figure 4.** Tangleogram highlighting the corresponde[illegible] one positive participant.

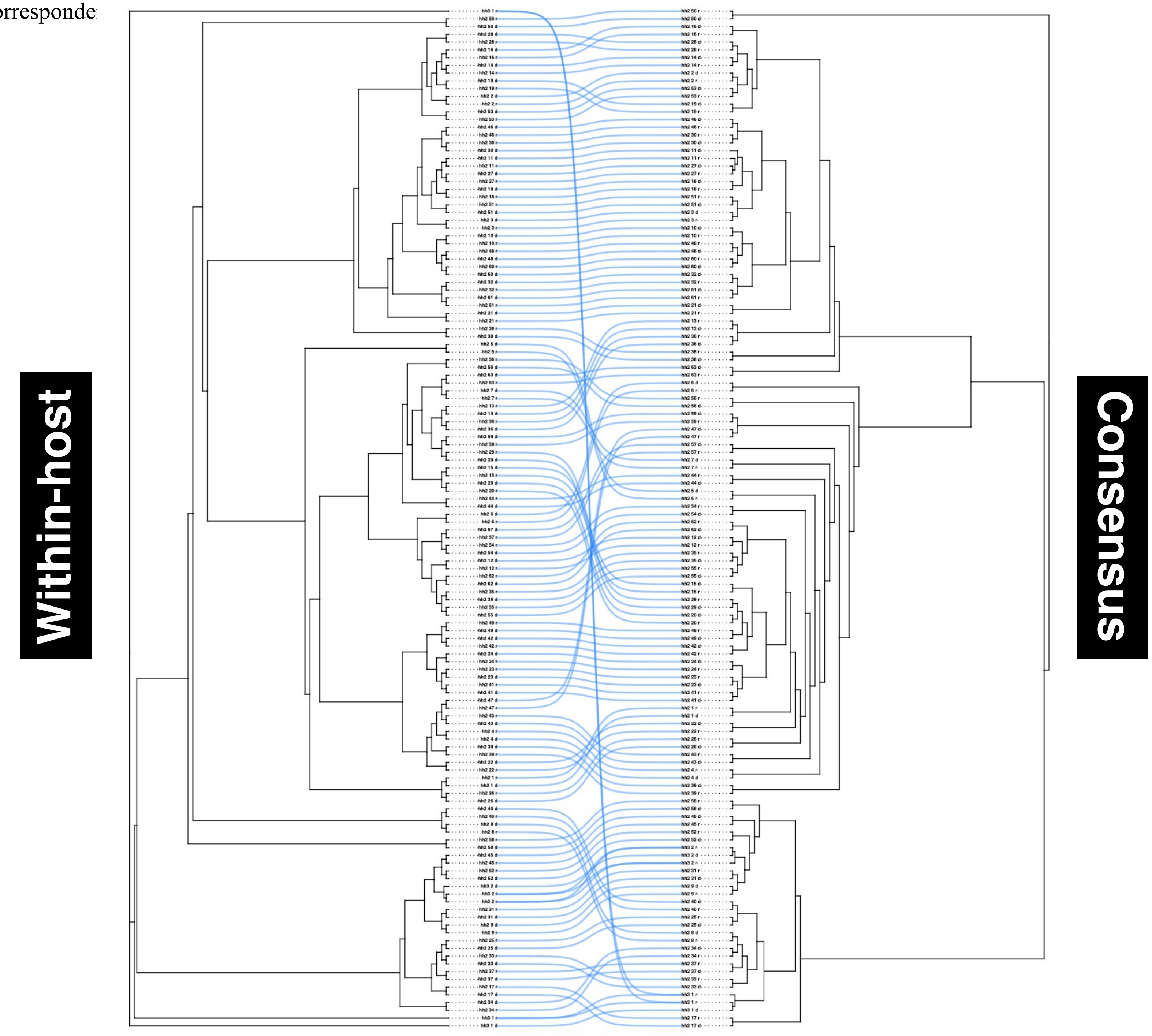

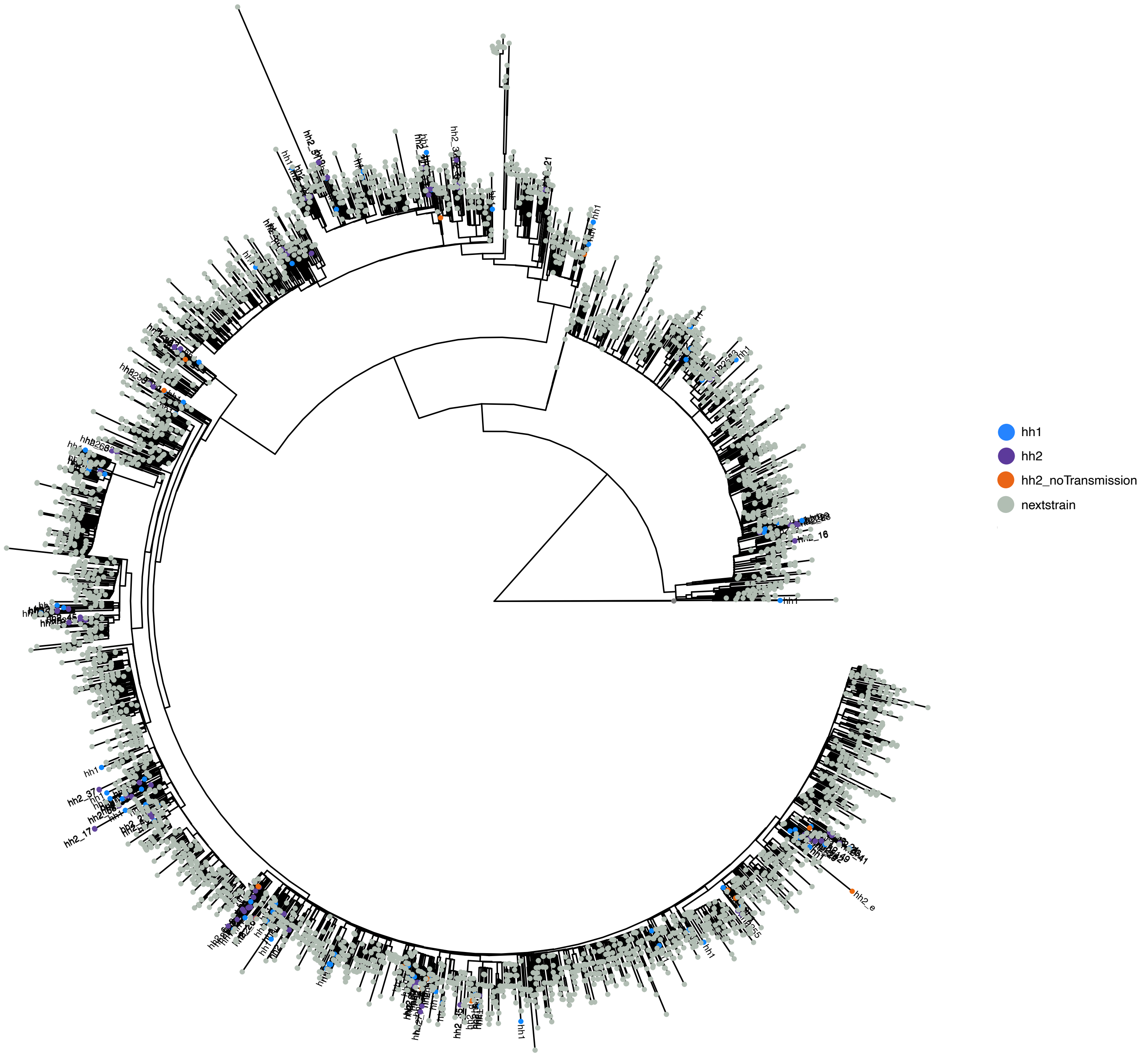


**Supplementary Figure 5.** Maximum likelihood phylogenetic tree generated using 4219 high coverage consensus sequence together with the genomic Virus Watch cohort. Turquoise dots indicate households (HH) with only one individual positive, purple dots refer to HH with more than one participant positive, orange dots highlight HH with no direct transmission, and in gray are the Nexstrain consensus sequences.

**Supplementary Figure 6.** Posterior probability of direct transmission for all households with more than one positive participant. Y axis represent infectors (who infected whom) and x axis infectees (individuals who were infected). Data plotted per lineage.

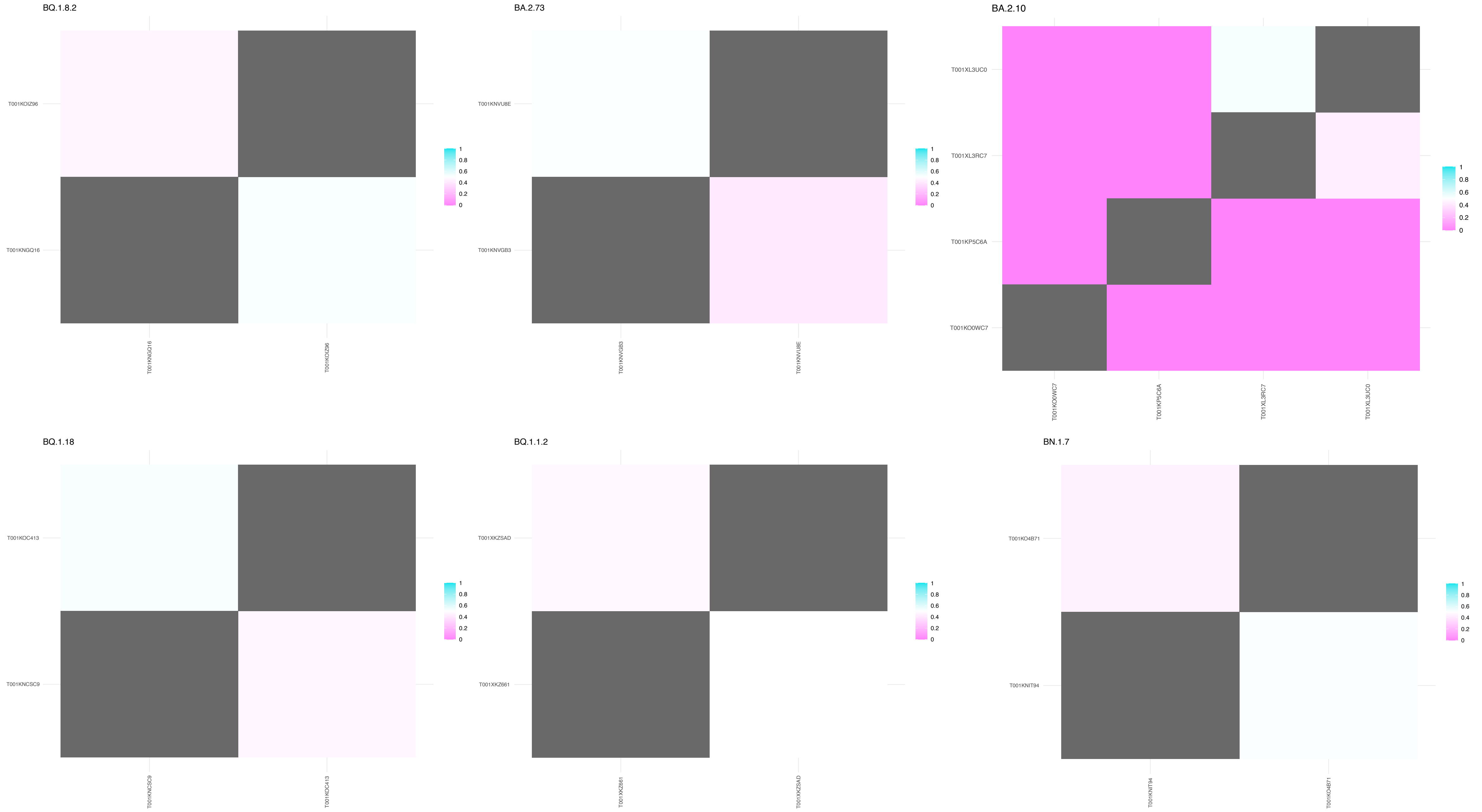

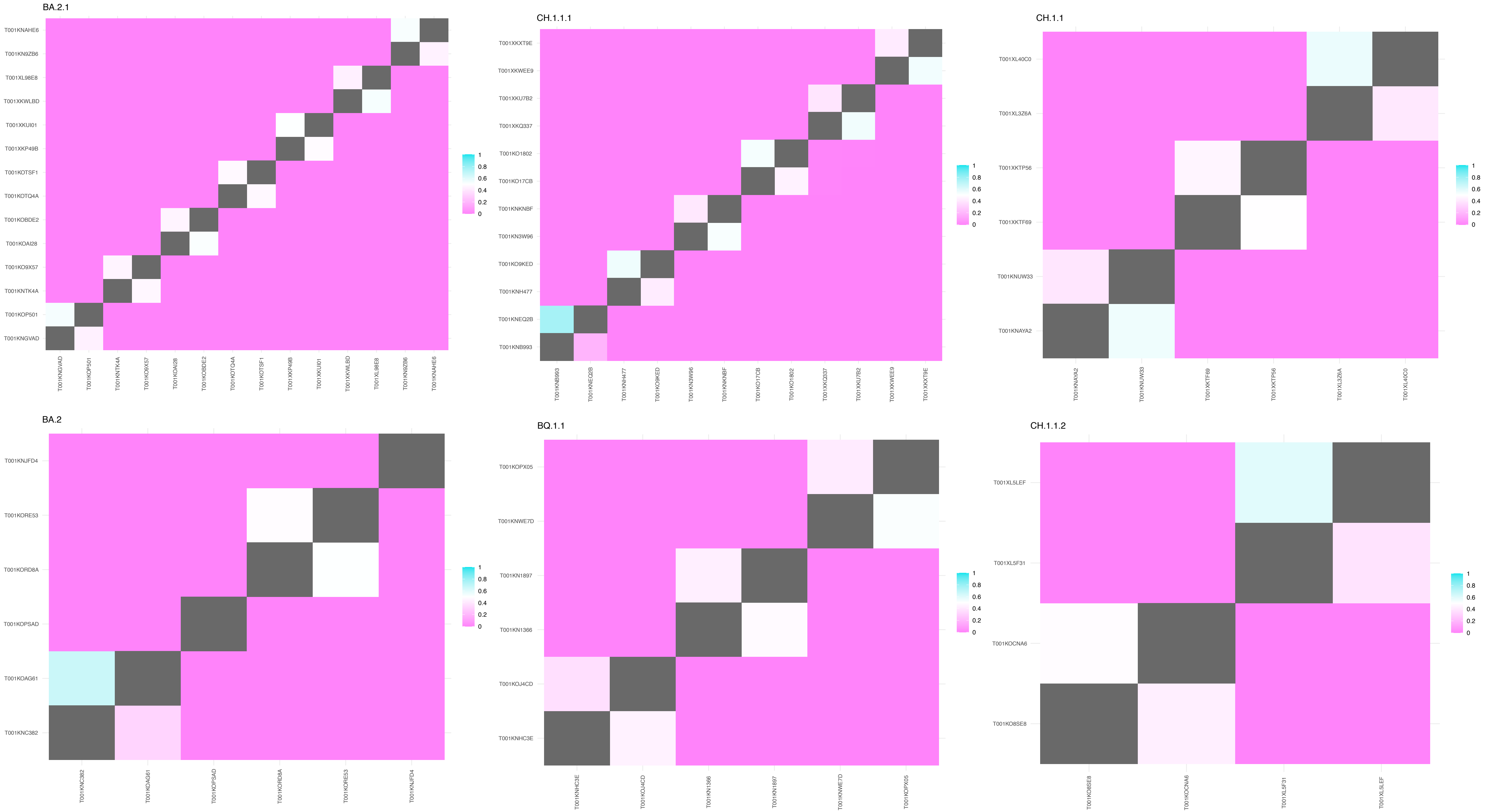
BA.2.1
T001KNAHE6
T001KN9ZB6
T001XL98E8
T001XKWLBD
T001XKUI01
T001XKP49B
T001KOTSF1
T001KOTQ4A
T001KOBDE2
T001KOAI28
T001KO9X57
T001KNTK4A
T001KOP501
T001KNGVAD
1
0.8
0.6
0.4
0.2
0
CH.1.1.1
T001XKXT9E
T001XKWEE9
T001XKU7B2
T001XKQ337
T001KO1802
T001KO17CB
T001KNKNBF
T001KN3W96
T001KO9KED
T001KNH477
T001KNEQ2B
T001KNB993
CH.1.1
T001XL40C0
T001XL3Z6A
T001XKTP56
T001XKTF69
T001KNUW33
T001KNAYA2
BA.2
T001KNJFD4
T001KORE53
T001KORD8A
T001KOPSAD
T001KOAG61
T001KNC382
BQ.1.1
T001KOPX05
T001KNWE7D
T001KN1897
T001KN1366
T001KOJ4CD
T001KNHC3E
CH.1.1.2
T001XL5LEF
T001XL5F31
T001KOCNA6
T001KO8SE8

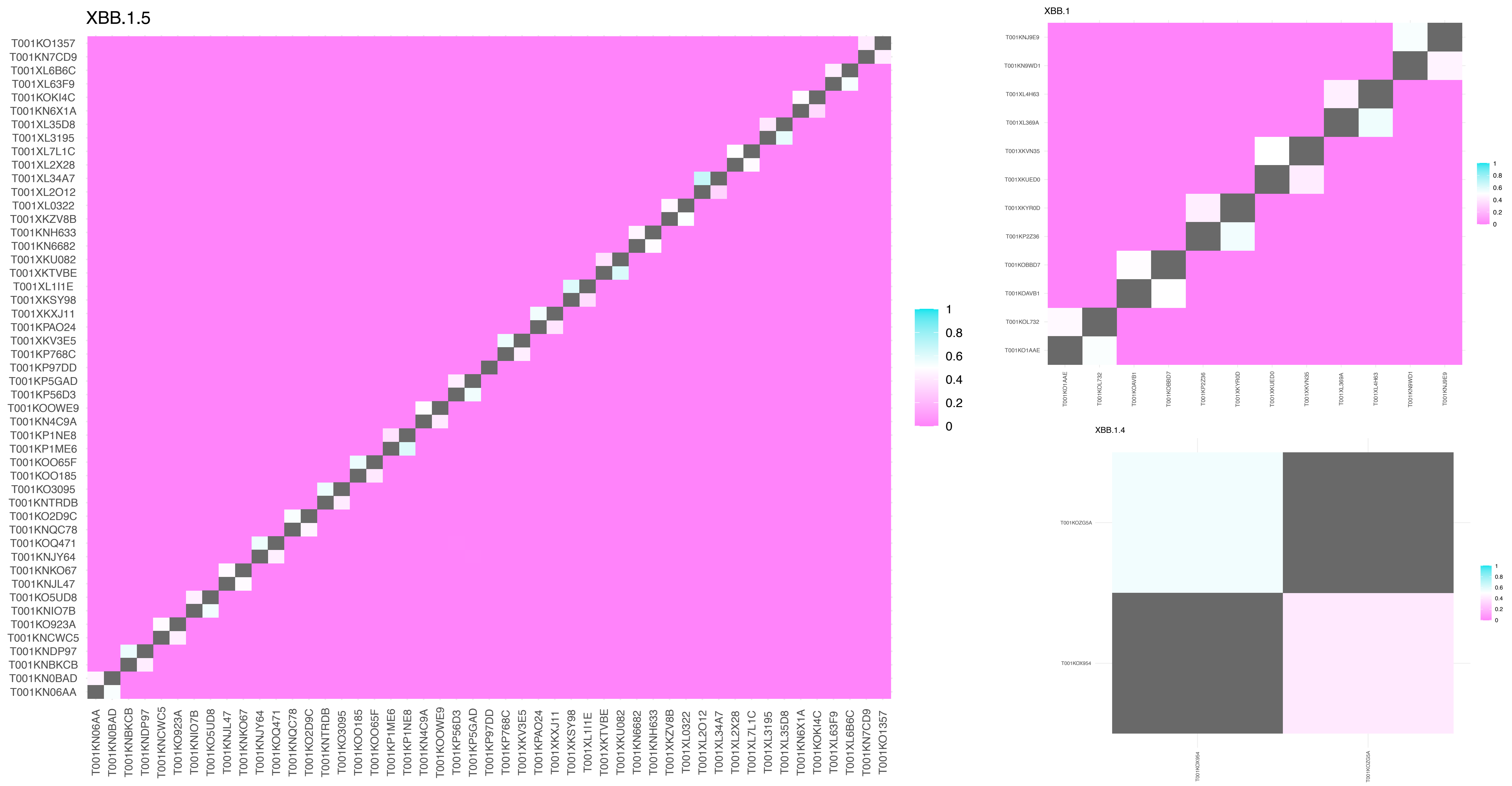
XBB.1.5
T001KO1357
T001KN7CD9
T001XL6B6C
T001XL63F9
T001KOKI4C
T001KN6X1A
T001XL35D8
T001XL3195
T001XL7L1C
T001XL2X28
T001XL34A7
T001XL2O12
T001XL0322
T001XKZV8B
T001KNH633
T001KN6682
T001XKU082
T001XKTVBE
T001XL1I1E
T001XKSY98
T001XKXJ11
T001KPAO24
T001XKV3E5
T001KP768C
T001KP97DD
T001KP5GAD
T001KP56D3
T001KOOWE9
T001KN4C9A
T001KP1NE8
T001KP1ME6
T001KOO65F
T001KOO185
T001KO3095
T001KNTRDB
T001KO2D9C
T001KNQC78
T001KOQ471
T001KNJY64
T001KNKO67
T001KNJL47
T001KO5UD8
T001KNIO7B
T001KO923A
T001KNCWC5
T001KNDP97
T001KNBKCB
T001KN0BAD
T001KN06AA
T001KN06AA
T001KN0BAD
T001KNBKCB
T001KNDP97
T001KNCWC5
T001KO923A
T001KNIO7B
T001KO5UD8
T001KNJL47
T001KNKO67
T001KNJY64
T001KOQ471
T001KNQC78
T001KO2D9C
T001KNTRDB
T001KO3095
T001KOO185
T001KOO65F
T001KP1ME6
T001KP1NE8
T001KN4C9A
T001KOOWE9
T001KP56D3
T001KP5GAD
T001KP97DD
T001KP768C
T001XKV3E5
T001KPAO24
T001XKXJ11
T001XKSY98
T001XL1I1E
T001XKTVBE
T001XKU082
T001KN6682
T001KNH633
T001XKZV8B
T001XL0322
T001XL2O12
T001XL34A7
T001XL2X28
T001XL7L1C
T001XL3195
T001XL35D8
T001KN6X1A
T001KOKI4C
T001XL63F9
T001XL6B6C
T001KN7CD9
T001KO1357
1
0.8
0.6
0.4
0.2
0
XBB.1
T001KNJ9E9
T001KN9WD1
T001XL4H63
T001XL369A
T001XKVN35
T001XKUED0
T001XKYR0D
T001KP2Z36
T001KOBBD7
T001KOAVB1
T001KOL732
T001KO1AAE
T001KO1AAE
T001KOL732
T001KOAVB1
T001KOBBD7
T001KP2Z36
T001XKYR0D
T001XKUED0
T001XKVN35
T001XL369A
T001XL4H63
T001KN9WD1
T001KNJ9E9
1
0.8
0.6
0.4
0.2
0
XBB.1.4
T001KOZG5A
T001KOX954
T001KOX954
T001KOZG5A
1
0.8
0.6
0.4
0.2
0

**Supplementary Figure 7.** Total number of shared variants: inferred transmission pairs *vs* random pairs. Allele frequency (AF) considered were 5-95% in blue and 10-95% in pink. a) random pairs are shuffled from households with only one positive participant; b) random pairs are shuffled from the inferred transmission pairs.

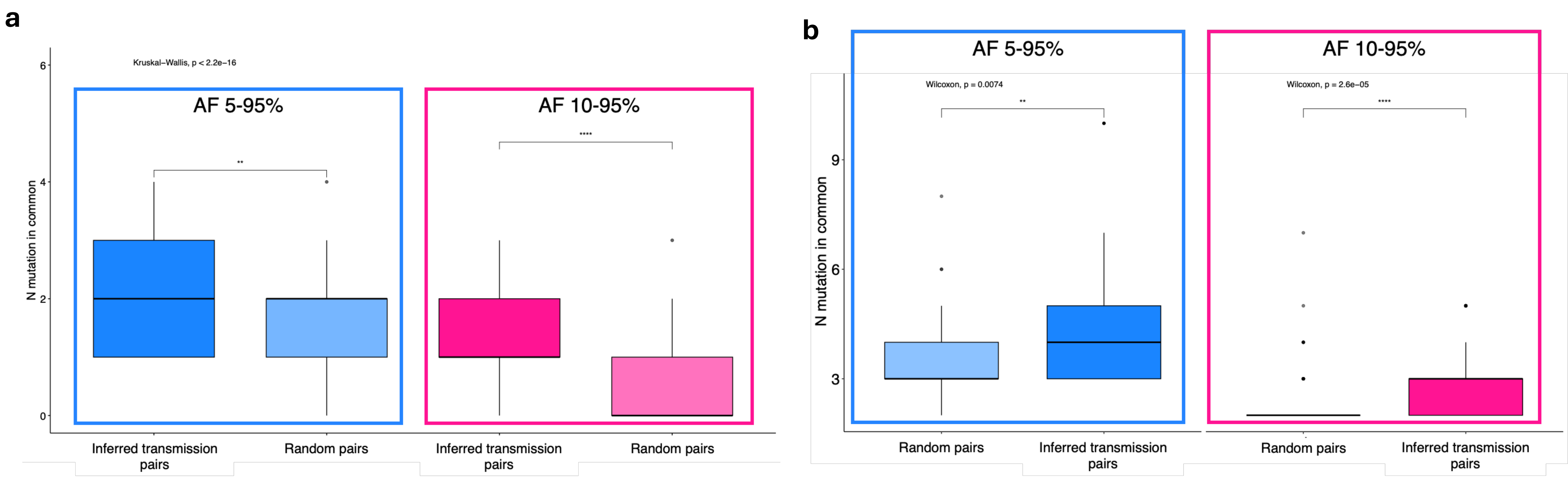
